# Replication of a GWAS signal near *HLA-DQA2* with acute myeloid leukemia using a disease-only cohort and external population-based controls

**DOI:** 10.1101/2024.09.26.24314422

**Authors:** Rose Laflamme, Véronique Lisi, Josée Hébert, Guy Sauvageau, Sébastien Lemieux, Vincent-Philippe Lavallée, Guillaume Lettre

## Abstract

Acute myeloid leukemia (AML) is the most common type of acute leukemia in adults. Its risk factors include rare and highly penetrant somatic mutations. Genome-wide association studies (GWAS) have also identified four common inherited variants associated with AML risk, but these findings have not yet been confirmed in many independent datasets. Here, we performed a replication study with 567 AML cases from the Leucegene cohort and 1,865 controls from the population-based cohort CARTaGENE (CaG). Because genotypes were generated using different technologies in the two datasets (e.g. low- vs. high-coverage whole-genome sequencing), we applied stringent quality-control filters to minimize type I errors. We showed using data reduction methods (e.g. principal component analysis [PCA] and uniform manifold approximation and projection [UMAP]) that our approach successfully integrated the Leucegene and CaG genetic data. We replicated the association between cytogenetically normal (CN)-AML and rs3916765, a variant located near *HLA-DQA2* (odds ratio [95% confidence interval] = 1.88 [1.21-2.93], P-value=0.005). The effect size of this association was stronger when we restricted the analyses to AML patients with *NPM1* mutations (odds ratios >2.35). We found *HLA-DOB* to be the most significantly upregulated gene in Leucegene participants with the CN-AML protective A-allele at rs3916765. We further found that several HLA class II genes are also differentially expressed albeit at lower statistical significance. Our results confirm that a common genetic variant at the HLA locus associates with AML risk, providing new opportunities to improve disease prognosis and treatment.

## INTRODUCTION

Acute myeloid leukemia (AML) is characterized by the dysregulated clonal expansion of malignant myeloblasts in the bone marrow (1). Although it is one of the most common leukemias in adults, it represents only 1% of all cancers (1). In 2019, 1,160 Canadians were diagnosed with AML (2). AML is highly lethal in older adults and very aggressive in younger individuals, with a 5-year relative survival rate of ∼32% (3). It is a genetically complex disease with distinct AML subgroups defined by specific genetic abnormalities (4). Different classes of germline genetic variation can influence the risk to develop AML, including monogenic predispositions, as well as more common genetic variants with individually modest impact on AML risk (4,5). While the best-established associations with AML are monogenic in nature, recent genome-wide association studies (GWAS) have identified four susceptibility loci for AML, including variants in or near *KMT5B*, *IRF4*, *HLA-DQA2*, and *BICRA* (1,6–8). The variant near *HLA-DQA2* is associated with cytogenetically normal AML (CN-AML) (6). Additional replication in independent AML case-control cohorts would reinforce the evidence that these common, modest effect variants are *bona fide* AML risk factors.

One challenge when working with an uncommon disease like AML is to have a sufficiently large sample size to enable the discovery or replication of weak-to-moderate effect size genetic variants. Given a fixed budget, one approach consists in sequencing (or genotyping) the genome of AML patients only, and to use publicly available non-affected cohorts as control samples. However, combining genetic data from cases and controls that were generated using a different approach or at a different time represents a difficult analytical task because it can result in technical batch effects that can lead to an increase type I error rate (9–11).

We performed a replication study of AML susceptibility GWAS loci using genetic data from an AML cohort and an external set of controls. Specifically, we used 567 patients of European ancestry from the Leucegene cohort as AML cases (12). These patients were recruited in different centers from Quebec (Canada) and have low-pass whole-genome sequence (WGS) data available (mean coverage 3.25X) of either a tumoral sample, or both a tumoral and a normal sample. Because many of the Leucegene participants are French Canadians, a founder population (13), we selected controls from the CARTaGENE (CaG) population-based cohort. CaG includes ∼43,000 participants recruited between the ages of 40 and 69 from several regions of Quebec, including Greater Montreal (14). There are currently 2,173 CaG participants with high-coverage WGS data available (mean coverage >30X), including 1,756 French Canadians, as well as 163 and 131 individuals with four grandparents born in Haiti and Morocco, respectively. We merged the Leucegene and CaG data to replicate the association between AML risk and the known GWAS AML variants. In exploratory analyses, we also analyzed the association between AML and polygenic risk scores (PRS) calculated using variants from leukemia or blood-cell traits GWAS.

## RESULTS

### Integration of Leucegene AML cases and CaG controls

To call genotypes in the Leucegene cohort, we used GLIMPSE and phased haplotypes from the 1000 Genomes Project (**Methods**) (15,16). We applied stringent filters to remove poorly imputed variants and samples with low-quality sequence data to obtain a dataset that totals 16,600,819 variants and 598 samples (**Supplemental Fig. 1** and **Methods**). To validate the GLIMPSE variant calls, we calculated the genotype concordance rate with whole-exome sequence (WES) variant calls available from 336 Leucegene participants: across the four WES sequencing batches, concordance was high (>97%) (**Methods**), suggesting that we can accurately call genotypes from low-pas WGS data.

To avoid false positive associations due to technical differences between the Leucegene and CaG datasets, we applied additional filters before merging the genotype data. We restricted downstream analyses to individuals of European ancestry (567 AML cases and 1,865 controls) and to 7,658,313 common variants (minor allele frequency [MAF] in the combined dataset ≥1%) with consistent allele frequencies in the gnomAD Non-Finnish European population (**Supplemental Fig. 2** and **Methods**) (17). After merging the Leucegene and CaG datasets, we applied data reduction methods – principal component analysis (PCA) and unified manifold approximation and projection (UMAP) - to visualize potential population stratification (18). First, we confirmed that the Leucegene and CaG participants cluster with European-ancestry individuals from the 1000 Genomes Project (**Fig. 1A-B**). Second, we repeated these analyses with only the Leucegene and CaG participants: both cohorts are well-integrated together, except for a cluster of 55 individuals (54 AML cases and one CaG control) that cluster separately from the main population (**Fig. 1C-D**). Additional analyses using the different sub-populations from the 1000 Genomes Project’s European superpopulation showed that these 55 individuals clustered with the Tuscan (TSI) samples, suggesting that they might be Quebec residents of Italian descent (**Supplemental Fig. 3**).

**Figure 1.**
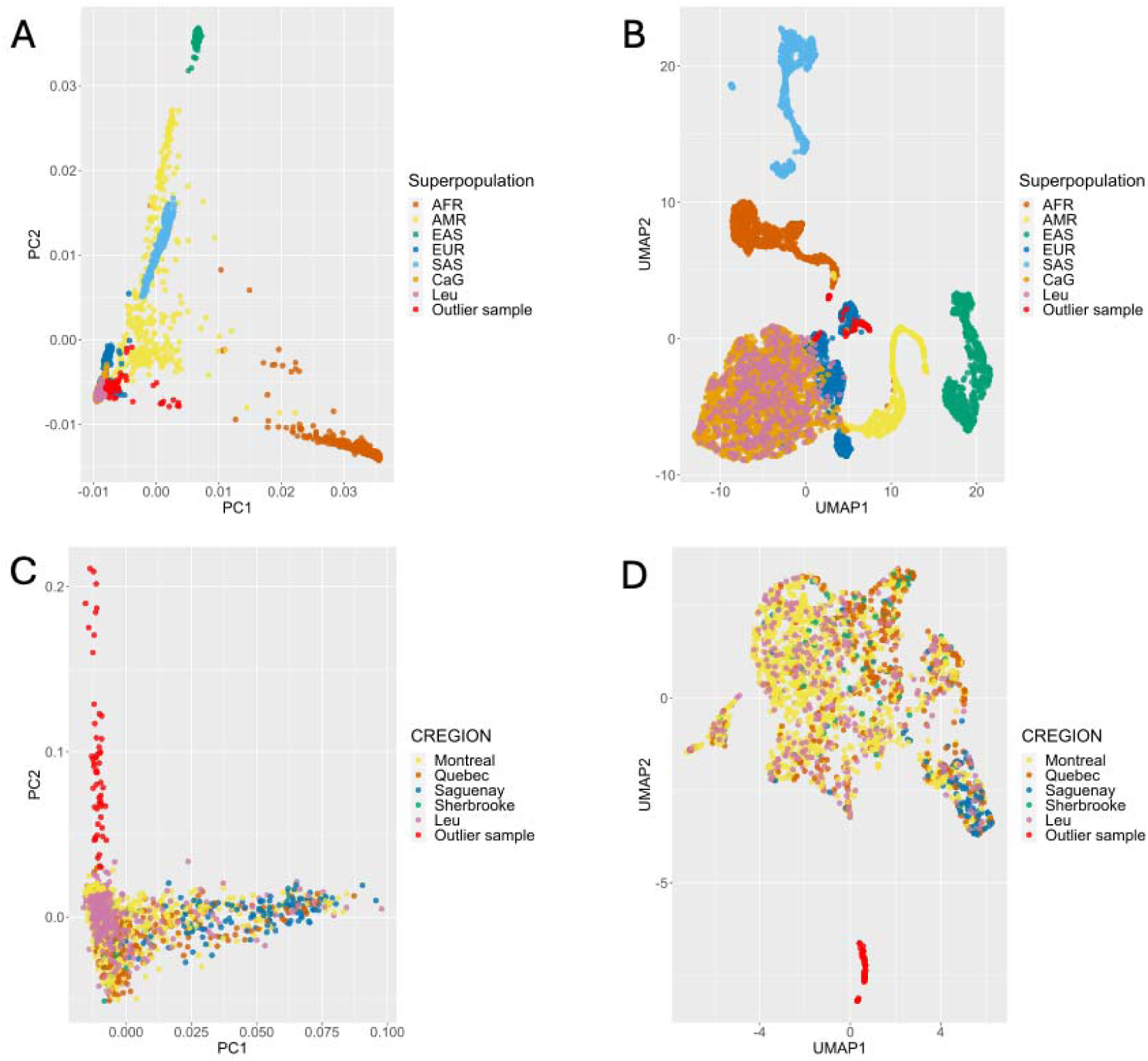
Genetic ancestry in the acute myeloid leukemia (AML) Leucegene and CARTaGENE (CaG) cohorts. Principal component analyses (PCA) (**A**,**C**) and uniform manifold approximation and projection (UMAP) representations (**B**,**D**) of the Leucegene and CaG participants, with (**A-B**) or without (**C-D**) individuals from the 1000 Genomes Project. Superpopulation refers to the 1000 Genomes Project labels (AFR=African, AMR=American, EAS=East Asian, EUR=European, SAS=South Asian). CREGION reffers to the region of recruitment for the CaG participants (Leu for AML patients). In red are outlier Leucegene and CaG samples that cluster with the Italian samples (TSI) from the 1000 Genomes Project (see also **Supplementary Fig. 3**).

### Replication of variants associated with AML risk

Lin et al. reported two genome-wide significant susceptibility loci for AML: rs4930561 in *KMT5B* associated with all AML types, and rs3916765 near *HLA-DQA2* associated with CN-AML (6). In our Leucegene-CaG dataset, we replicated the association between *HLA-DQA2*-rs3916765 and CN-AML (G-allele odds ratio (OR)=1.88, P-value=0.0052) (**Table 1**). This association between rs3916765 and CN-AML remained significant after excluding the 55 outlier samples described above (P-value=0.0076). The AML variant in *KMT5B* did not replicate in our study (**Table 1**). Two more variants have been reported to be associated with AML: rs75797233 near *BICRA* and rs12203592 in an intron of *IRF4* (7,8). rs75797233 was not imputed by GLIMPSE, but this can be due to its low minor allele frequency ([MAF=0.02]) (8). rs12203592 was imputed by GLIMPSE but was excluded because it did not meet our imputation quality threshold (imputation R^2^=0.898).

**Table 1.** Replication results in the Leucegene-CARTaGENE dataset for two variants previously associated with acute myeloid leukemia (AML) by genome-wide association study (GWAS). OR [95% CI], odds ratio and 95% confidence interval; NFE, non-Finnish Europeans.

| Variant | Risk allele | Gene | All AML<br>N=567 cases, 1865 controls |  | CN-AML<br>N=239 cases, 1865 controls |  | Allele frequency of the risk allele |  |  |  |
| --- | --- | --- | --- | --- | --- | --- | --- | --- | --- | --- |
|  |  |  | P-value | OR [95% CI] | P-value | OR [95% CI] | gnomAD NFE | Control | Cases all AML | Cases CN-AML |
| 6:32717773<br>G>A<br>rs3916765 | G | *HLA-DQA2* | 0.13 | 1.22<br>[0.94-1.58] | 0.005 | 1.88<br>[1.21-2.93] | 0.89 | 0.90 | 0.92 | 0.95 |
| 11:68164294<br>G>A<br>rs4930561 | A | *KMT5B* | 0.10 | 1.13<br>[0.98-1.30] | 0.067 | 1.21<br>[0.99-1.48] | 0.50 | 0.49 | 0.52 | 0.54 |

AML are classified into molecular subgroups defined by genetic abnormalities (4). AML with mutations at the C-terminus of the protein encoded by the nucleophosmin gene *NPM1* on chromosome 5 represents a third of AML cases, and 60% of CN-AML cases (4,19). In the Leucegene-CaG dataset, when we stratified the analyses to AML cases with *NPM1* mutations (N*_NPM1_*=177 cases) or with *NPM1* mutations and a normal karyotype (N_CN-*NPM1*_=145 cases), the association with HLA-DQA2- rs3916765 was stronger (with respective OR of 2.37 and 2.70 for the G-allele) (**Supplemental Table 1**).

### Expression quantitative trait locus (eQTL) analysis at rs3916765

We queried the GTEx whole-blood eQTL dataset and found that genotypes at rs3916765 are associated with the expression of *HLA-DOB* and *HLA-DQA2* (**Fig. 2A**). We then investigated in the Leucegene RNA-sequencing dataset (N=567) if rs3916765 was also an eQTL (20). Differential gene expression analysis between participants with the protective allele (AA or GA) and those without (GG) identified 360 differentially expressed genes at nominal significance (P-value <0.05, **Fig. 2B**). Of those, the most significantly differentially expressed gene was *HLA-DOB* (adjusted P-value=4.9×10^-23^). *HLA-DQA2* was the second most differentially expressed class II HLA gene (nominal P-value=0.0079), although it did not reach significance after multiple testing correction. Globally, most of the class II HLA genes were differentially expressed between participants with and without the protective A-allele at rs3916765 (**Fig. 2B-C**). The observed differential expression was independent of the *NPM1* mutational status since *HLA-DOB* was significantly differentially expressed (after multiple testing correction) regardless of the *NPM1* genotype (**Supplemental Figure 4**).

**Figure 2.**
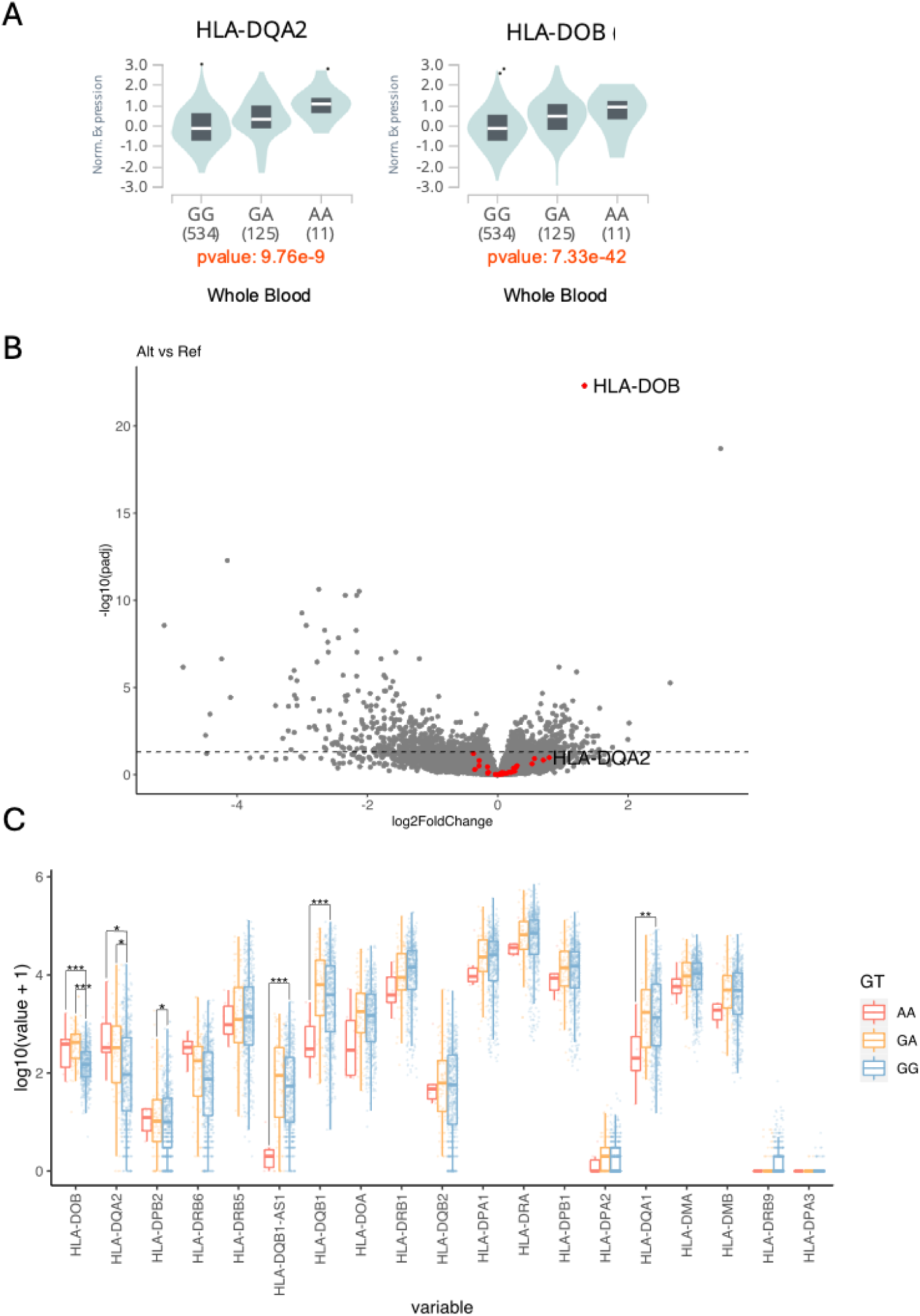
rs3916765 is an expression quantitative trait locus (eQTL) for HLA class II genes in whole-blood. (**A**) rs3916765 genotype-specific expression of *HLA-DQA2* and *HLA-DOB* in whole-blood from GTEx. (**B**) Differentially expressed genes between Leucegene participants with reference genotype (GG, N=487) and non-reference genotype (GA and AA, N=80) at rs3916765. Genes in red are HLA class II genes. (**C)** Genotype-specific expression of HLA class II genes at rs3916765 in the Leucegene participants. *P-value <0.05; **P-value <0.01; ***P-value <0.001.

### Polygenic risk scores (PRS) for leukemia and blood cell traits

There are currently no published PRS for AML. As exploratory analyses, we calculated PRS in the Leucegene-CaG dataset for acute lymphocytic leukemia (ALL) and chronic lymphocytic leukemia (CLL). Since AML is a malignancy of the myeloid lineage, we also calculated PRS for seven blood-cell traits of the myeloid lineage: basophil count, eosinophil count, monocyte count, neutrophil count, platelet count, red blood cell count, and total white blood cell count. When we tested the association between AML risk and the different PRS, we observed no significant associations after correcting for the number of tests performed (**Fig. 3** and **Supplemental Fig. 5**). The most significant association was between AML and the neutrophil PRS: the risk of AML increased by 11.8% for each standard deviation increased in the neutrophil PRS (nominal P-value=0.021). Except for two, the variants used in the calculation of the PRS are not significantly associated with AML nor CN-AML when taken individually (**Supplemental Fig. 6**).

**Figure 3.**
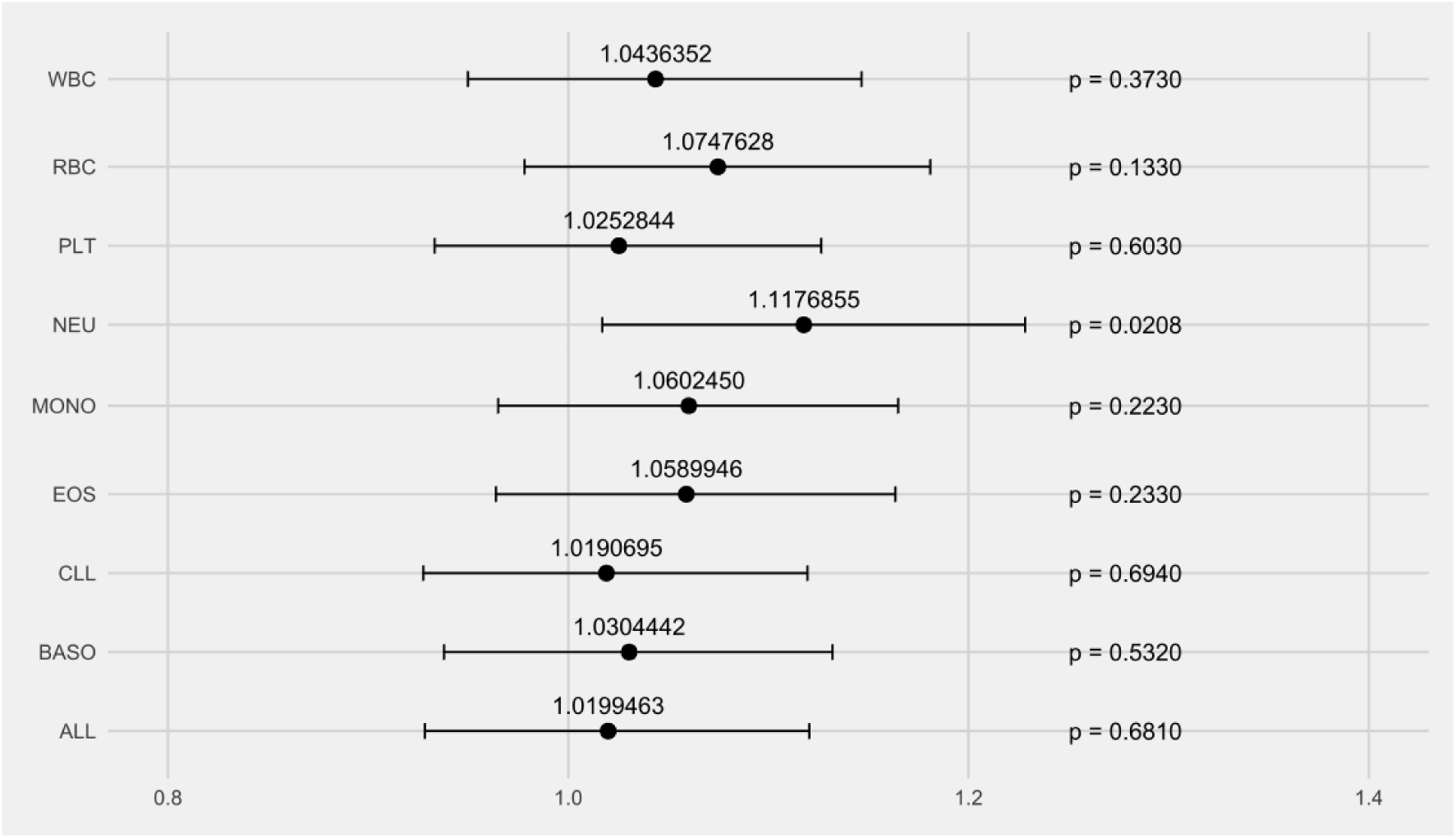
Association of leukemia and blood-cell traits polygenic risk scores (PRS) with acute myeloid leukemia (AML) risk. We calculated PRS for acute lymphocytic leukemia (ALL), chronic lymphocytic leukemia (CLL), and seven blood-cell traits in the Leucegene AML cases and the CARTaGENE controls. The black dots correspond to the odds ratios per one standard deviation in the PRS. The whiskers are the 95% confidence intervals for the odds ratios. WBC, white blood cell count; RBC, red blood cell count, PLT platelet count, NEU, neutrophil count; MONO, monocyte count; EOS, eosinophil count; BASO, basophil count.

## DISCUSSION

AML is one of the most common forms of leukemia affecting adults. Its genetic etiology is complex, involving well-established monogenic alterations as well as modest effect and common germline variants (1,4,5). GWAS have started to describe the polygenic architecture of AML (1,6–8), but efforts have remained limited. To strengthen previous AML GWAS findings, we performed a replication study using data from the well-characterized Leucegene AML cohort and controls from the population-matched CaG cohort. Despite our limited sample size, we could replicate the association between CN-AML and genotypes at *HLA-DQA2*-rs3916765. The effect size of this association was strong for a common variant (OR=1.88), and even stronger when restricting the analysis to patients with an *NPM1* mutation (OR=2.37) (**Table 1** and **Supplemental Table 1**). The risk G-allele at rs3916765 has a frequency of 0.90 in CaG controls and of 0.92 in AML patients, but reaches 0.95 in CN-AML patients and 0.96 in *NPM1*-mutated AML patients or CN-*NPM1*-mutated AML patients. Mutant NPM1 proteins can contain peptides that bind HLA class I and II molecules more efficiently, making AML cells more vulnerable to immune recognition (21). Genotypes at rs3916765 might affect gene expression program of antigen presenting cells, thereby impacting their role in the immune response by providing a protective effect. This could explain that DNA sequence variants near *HLA-DQA2* have stronger effect sizes in *NPM1*-mutated AML or CN-*NPM1*-mutated AML. However, our eQTL analyses indicate that genotypes at rs3196765 associate with *HLA-DOB* and *HLA-DQA2* expression independently of *NPM1* mutations, suggesting that the gene regulatory effect is not dependent on *NPM1* status. Given the complexity of the HLA locus in the human genome, more functional work is needed to determine how genotypes at rs3916765 affects AML risk in combination with the *NPM1* mutation status.

To perform our analyses, we faced two important challenges. First, we called genotypes from low-pass WGS in the Leucegene cohort. While there are limitations to this approach (e.g. the quality for rare variants), GLIMPSE generated high-quality genotype calls when compared to genotypes obtained using high-coverage WES. Second, because our AML cases and controls were profiled on different platforms and at different times, any technical artifacts (or batch effects) may lead to spurious associations. To minimize this risk, we applied very stringent QC thresholds on the datasets. We explicitly decided not to perform genome-wide association testing, but to focus on the replication of known AML variants. We also reasoned that PRS would be less sensitive to this issue, as a false positive result would imply systematic biases across 100s of variants.

In conclusion, the HLA locus is a validated and relatively strong risk factor for AML. Whether it impacts response to treatment is currently unknown. More generally, our study provides bioinformatic guidance for future genetic efforts that require combining different datasets generated using different methods. We also validated CaG as an external cohort of controls to enable association studies in the Quebec population.

## METHODS

### Whole-genome (WGS) and whole-exome (WES) sequencing of Leucegene participants

WGS data of the cohort was first published and described here (22). Exome germline data was generated using Agilent’s SureSelect kit following manufacturer’s instruction. Libraries were sequenced on Illumina NovaSeq with paired-end 100.

### Genotype imputation using low-depth WGS data

Low-pass WGS (mean coverage of 3.25x) was available for 649 AML patients of the Leucegene cohort (https://data.leucegene.iric.ca/). Of those, both tumoral and normal WGS was performed in 334 patients, only tumoral WGS in 311 patients and only germline WGS in 4 patients. Sources of tumoral cells were either blood or bone marrow for tumor samples, and sources of normal cells were either saliva or buccal swabs. Since some patients possess a pair of samples, we combined their tumoral and normal sample bam files with samtools 1.17 (23) prior to genotype imputation. We recalibrated bases with GATK 4.2.5.0 BaseRecalibrator and ApplyBQSR (24). As a reference panel for genotype imputation with GLIMPSE 1.1.1 (15), we used the phased 1000G phase 3 hg38 data (available here: http://ftp.1000genomes.ebi.ac.uk/vol1/ftp/data_collections/1000G_2504_high_coverage/working/20201028_3202_phased/) (16). We used a Nextflow pipeline (https://github.com/CERC-Genomic-Medicine/glimpse_pipeline) that runs GATK HaplotypeCaller (24) to generate genotype likelihoods at reference sites before GLIMPSE imputation. We removed multiallelic variants from the reference panel with bcftools 1.11 (23), since GLIMPSE can only handle biallelic variants. From the call set, we removed monomorphic variants and kept variants with an imputation quality R^2^ ≥ 0.95. To validate our GLIMPSE variant calls, we looked at the genotypic concordance between GLIMPSE calls and the Leucegene whole-exome sequence (WES) dataset. 336 Leucegene patients had their exome sequenced in four sequencing batches of 117, 123, 70, and 26 patients. To identify variants in WES data, we mapped reads to the genome using BWA-MEM (25), performed a cleaning and recalibration step using GATK and called the mutants using Mutect2 (v. 4.1.3.0) (24). We compared each batch separately. We removed variants from the WES dataset with more than 5% or 10% genotype missingness. We used PLINK2 to measure the concordance (26,27). With the 5% genotype missingness filter, the genotype concordance was 99.15%, 98.7%, 98.0%, and 98.1% for the four batches, while it was 98.3%, 98.2%, 97.5%, and 97.1% with the 10% genotype missingness filter. We also looked at the genotype concordance of common variants (with minor allele frequency higher than 1% or 5%). For the sequencing batch with 123 patients, the genotype concordance was 97.9% for variants with a minor allele frequency (MAF) >1%, and 97.3% for variants with a MAF >5%.

We performed standard quality-control (QC) steps of the Leucegene dataset using PLINK2. Using variants in linkage equilibrium, we looked at the heterozygosity of our samples, the concordance between reported and inferred sex, cryptic relatedness (kinship), and applied data reduction methods (principal component analysis [PCA] and uniform manifold approximation and projection [UMAP]) to visualize the data. We did not filter on genotype or sample missingness since there are no missing genotypes from the genotype imputation with GLIMPSE. We removed five patients because their heterozygosity was too low (F < -0.4), six patients because the declared sex did not match the inferred sex (F ≥70 for males and F <70 for females), three patients that were another patient’s first-degree relative (kinship coefficient >0.177), and 37 patients that were duplicated (patients that were sequenced twice to see the progression of the tumor or because of a relapse, kinship coefficient >0.354). For each pair of duplicates, we kept the sample that was sequenced first. After QC, 598 patients were left for downstream analyses.

### Inferring the genetic ancestry of Leucegene participants

Since self-declared ethnicity information about Leucegene patients was not collected, we inferred their genetic ancestry. We first projected Leucegene patients on the 1000G phase 3 dataset (n=2,548) and used UMAP and PCA visualization to assign each patient to a 1000G superpopulation (European, African, American, East Asian, or South Asian). To confirm our assignment, we used these superpopulations as the self-identified ethnicity variable (sire) for HARE (Harmonizing genetic ancestry and self-identified race/ethnicity) (28), but no Leucegene individual had their superpopulation changed. As expected, most patients are of European ancestry (94.81%, 567 patients), while 10 patients are of American ancestry (1.67%), 11 patients of African ancestry (1.84%), eight patients of East Asian ancestry (1.34%), and two patients of South Asian ancestry (0.33%). For the projection, we extracted variants common to the 1000G and the Leucegene datasets with PLINK2 (26,27) and merged them with PLINK1.9 (27,29). We kept variants in linkage equilibrium for the calculation of the principal components.

### AML tumoral subgroups classification

Tumors were classified as being cytogenetically normal or not and having mutations in *NPM1* or not. The cytogenetic information about the tumors is provided by the Quebec Leukemia Cell Bank (BCLQ). *NPM1* mutations were identified as previously described here (30).

### Whole-genome sequencing of CARTaGENE (CaG) participants

2,184 CaG participants (including 1,889 European-ancestry participants) were selected for high-coverage whole-genome DNA sequencing. The sequencing took place at the Genome Quebec *Centre d’Expertise et de Services (CES)* on Illumina NovaSeq sequencers using PCR-free libraries and a paired-end (2×150 bp) protocol. We used the Illumina DRAGEN pipeline to call and genotypes single nucleotide variants and insertions-deletions. After quality-control steps, 2,173 individuals were kept for downstream analyses.

### Integration of the Leucegene and CARTaGENE (CaG) genetic datasets

The integration of the CaG and Leucegene datasets was done with PLINK (26,27,29). We first normalized the position of the variants with the hg38 reference in the datasets separately and set the reference allele to be the one present in the hg38 reference genome. To control for technical differences between the cohorts, we applied strict filters on the individual cohorts before the merge. We kept individuals of European ancestry and variants with a MAF ≥1%. We removed variants with an HWE P-value <1×10^-3^ and variants for which the distance (DIST) between the allele frequency (AF) in the respective cohort and in gnomAD Non-Finnish European (gnomAD NFE) was superior to 0.06 for Leucegene and 0.055 for CaG (DIST = |gnomAD_NFE AF - cohort AF| / √2; variants that were not found or that were monomorphic in gnomAD NFE were removed). Individuals with European ancestry in CaG were defined as individuals with self-declared “White European” ethnicity and with a projection, while individuals with European ancestry in Leucegene were defined using PCA and UMAP projections on the 1000 Genomes Project dataset and with HARE (above). After applying the strict filters, there were 8,274,789 variants in Leucegene (8,328,109 before filtering) and 9,319,078 variants in CaG (9,939,090 before filtering). We extracted the variants that were common to both datasets before combining the cohorts. After the merge, there were 7,658,313 variants, 567 AML cases from Leucegene and 1,865 controls from CaG.

### Case-control association tests

We used logistic regression in PLINK2 for association testing. We used age, sex, and the 10 first principal components as covariates. The quantitative covariates were rescaled with --covar-variance-standardize. For the CN-AML, *NPM1*-mutated AML, and CN-*NPM1*-mutated AML replication studies, the principal components were recalculated with the set of individuals included in the test. We defined statistical significance as α=0.025 (Bonferroni correction for two tested variants [0.05/2]).

### eQTL analysis

Transcriptome generation was described previously (20) and is available here: https://www.ncbi.nlm.nih.gov/geo/query/acc.cgi?acc=GSE232130. Reads were mapped to the genome using STAR (v2.7.1) (31)and quantified using RSEM (v1.3.2) (32). RSEM generated pseudocount were used for differential expression analysis. Genes whose pseudo-count sums was less than 10 over the whole cohort were removed. Differential expression was performed using DEseq2 version 1.38 with default parameters (33). The rs3916765 genotype-specific expression in whole-blood from GTEx (GTEx Analysis Release V8) was obtained from the GTEx Portal on 09/20/2024 (34).

### Polygenic risk scores (PRS)

PRS were calculated with PLINK2 on the merged dataset with strict filters. For the seven blood-cell traits (BASO for basophil count, EOS for eosinophil count, MONO for monocyte count, NEU for neutrophil count, PLT for platelet count, RBC blood cell count, and WBC for white blood cell count), we used the variants from Chen et al. (35) and their European-ancestry variant weights, including 150 variants for BASO, 346 for EOS, 394 for MONO, 352 for NEU, 553 for PLT, 448 for RBC, and 443 for WBC, of which 112, 279, 340, 288, 457, 384, and 365 were present in the merged dataset with strict filters, respectively. For the acute lymphocytic leukemia (ALL) and chronic lymphocytic leukemia (CLL) PRS, we used variants from Berndt et al. (36), including 15 variants for ALL and 43 for CLL, of which 12 and 32 were present in our merged dataset, respectively. All variants included in the PRS were lifted over from build hg19 to hg38 with the online LiftOver tool from the UCSC Genome Browser (37). We applied an inverse normal transformation to the PRS and did a logistic regression to test their association with AML, correcting for age, sex and the first ten principal components. We defined statistical significance as α=0.0056 (Bonferroni correction for nine tested PRS [0.05/9]).

## ACKNOWLEDGMENTS

The authors wish to thank Muriel Draoui for Leucegene project coordination and personnel at the Institute for Research in Immunology and Cancer (IRIC) genomics platform for sequencing. The authors acknowledge the invaluable contribution of Quebec Leukemia Cell Bank (BCLQ) members, and of IRIC bioinformatic platform member Patrick Gendron. We thank all CaG participants. We also thank Daniel Talium for helpful advices on the best strategy to call variants from low-pass WGS data.

## DATA AVAILABILITY

Accessing the CaG WGS data requires the submission of a request application; the procedure is explained at: https://cartagene.qc.ca/en/researchers/access-request.html. The code used to analyze the data and generate the figures is available upon request.

## AUTHOR CONTRIBUTIONS

Conceived and designed the analyses: R.L., V.-P.L. and G.L.; Collected the data: J.H., G.S., S.L., V.-P.L., and G.L.; Contributed data: J.H., G.S., S.L., V.-P. L., and G.L.; Performed analyses: R.L. and V.L.; Secured funding and supervised the work: J.H., G.S., V.-P.L. and G.L.; Wrote the manuscript: R.L., V.P.-L. and G.L., with contributions from all authors.

## ETHICS

The Montreal Heart Institute Ethics Committee gave ethical approval for this work (Project #2017-2247).

