## Supplementary Tables and Figures for "Replication of a GWAS signal near *HLA-DQA2* with acute myeloid leukemia using a disease-only cohort and external population-based controls"

**Supplemental Figure 1.** Distribution of the imputation quality score for the polymorphic variants imputed with GLIMPSE from the Leucegene low-pass whole-genome sequence (WGS) data. Polymorphic variants with an imputation $R^2$ <0.95 (red line at $R^2$ = 0.95) were filtered out. There are 16.6M variants with $R^2$ ≥0.95, 18.6M with $R^2$ ≥0.9, 19.9M with $R^2$ ≥0.85, and 20.8M with $R^2$ ≥0.8.

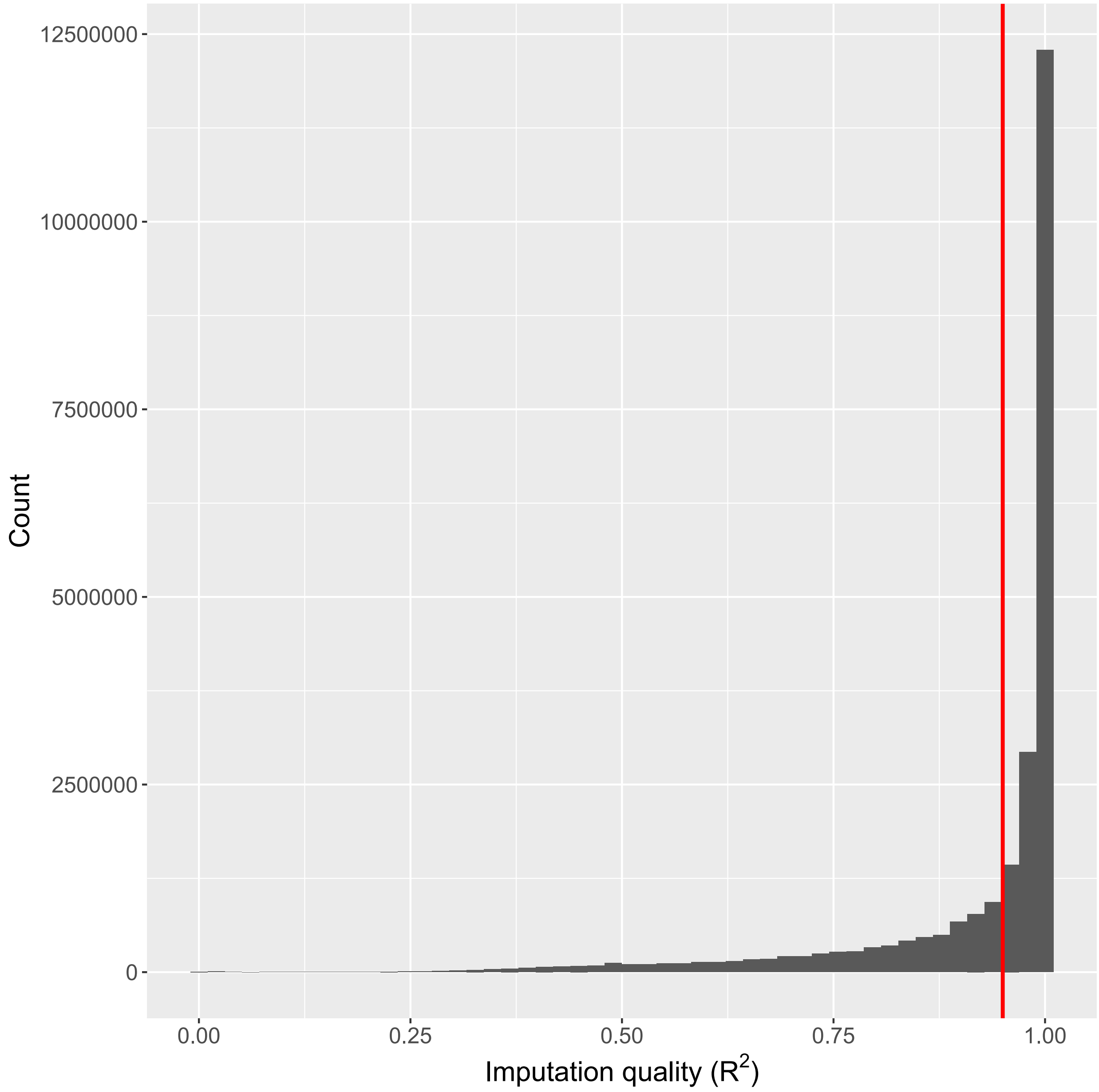

**Supplemental Figure 2.** Common variants (minor allele frequency ≥1%) that were excluded from the Leucegene (**A**) or CARTaGENE (CaG) (**B**) cohorts because they are out of the Hardy-Weinberg Equilibrium (HWE P-value <0.001, blue dots) or because their allele frequency is too distant from the same variant's allele frequency in gnomAD Non-Finnish Europeans (**Methods**, red dots). In **B**, variants in the first quadrant left to the diagonal consist mostly of variants with low genotype quality (more than 10% LowGQ at the participant level). **C** and **D** show allele frequency in Leucegene (x-axis) and CaG (y-axis) with or without excluded variants (in blue or red), respectively.

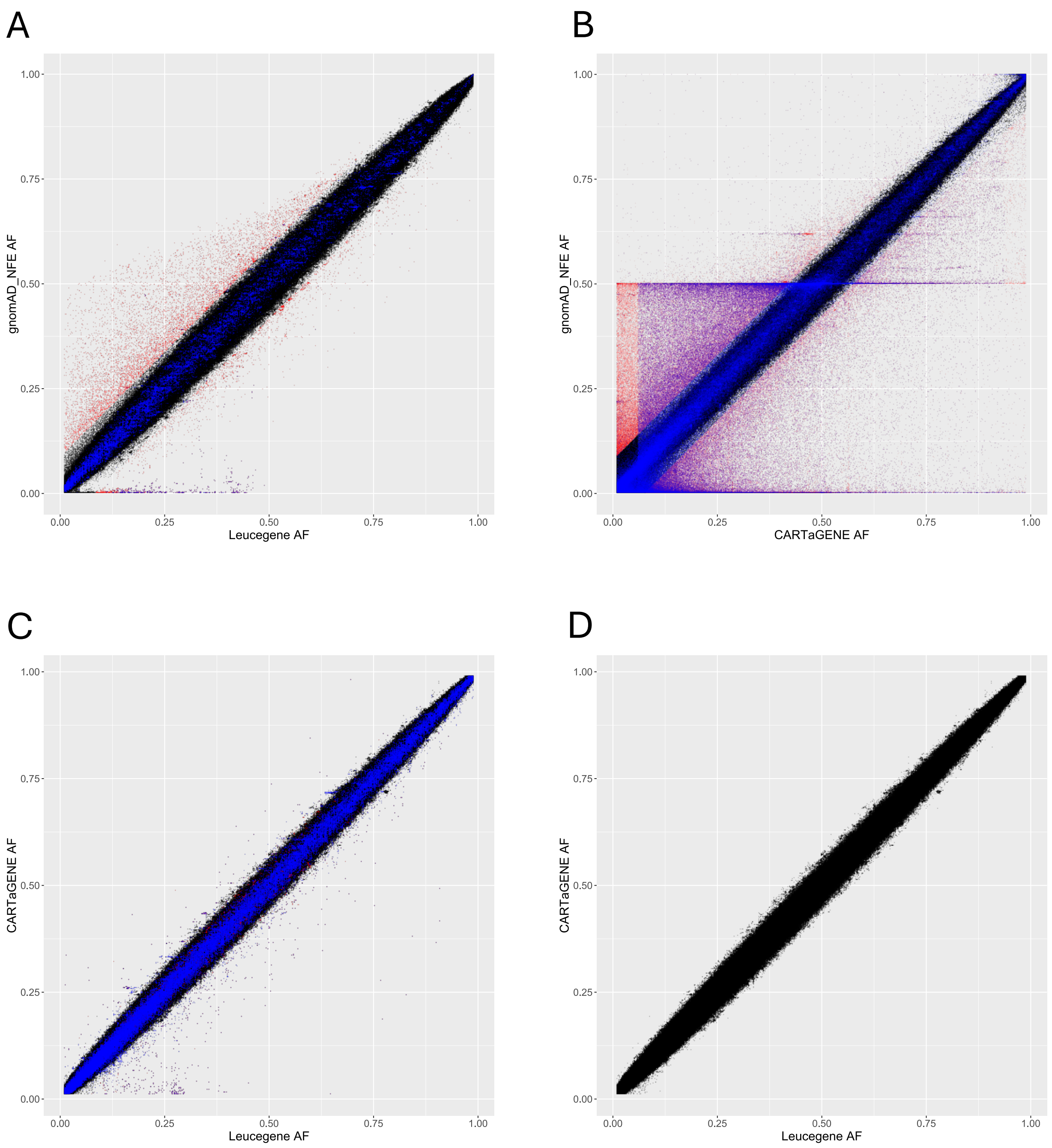

**Supplemental Figure 3.** Uniform manifold approximation and projection (UMAP) representation of the Leucegene and CARTaGENE (CaG) participants with European-ancestry participants from the 1000 Genomes Project. In red are outliers participants from Leucegene and CaG that mostly cluster with the Italian participants from the 1000 Genomes Project (CEU,CEPH; FIN,Finnish; GBR,British; IBS,Spanish; TSI,Tuscan).

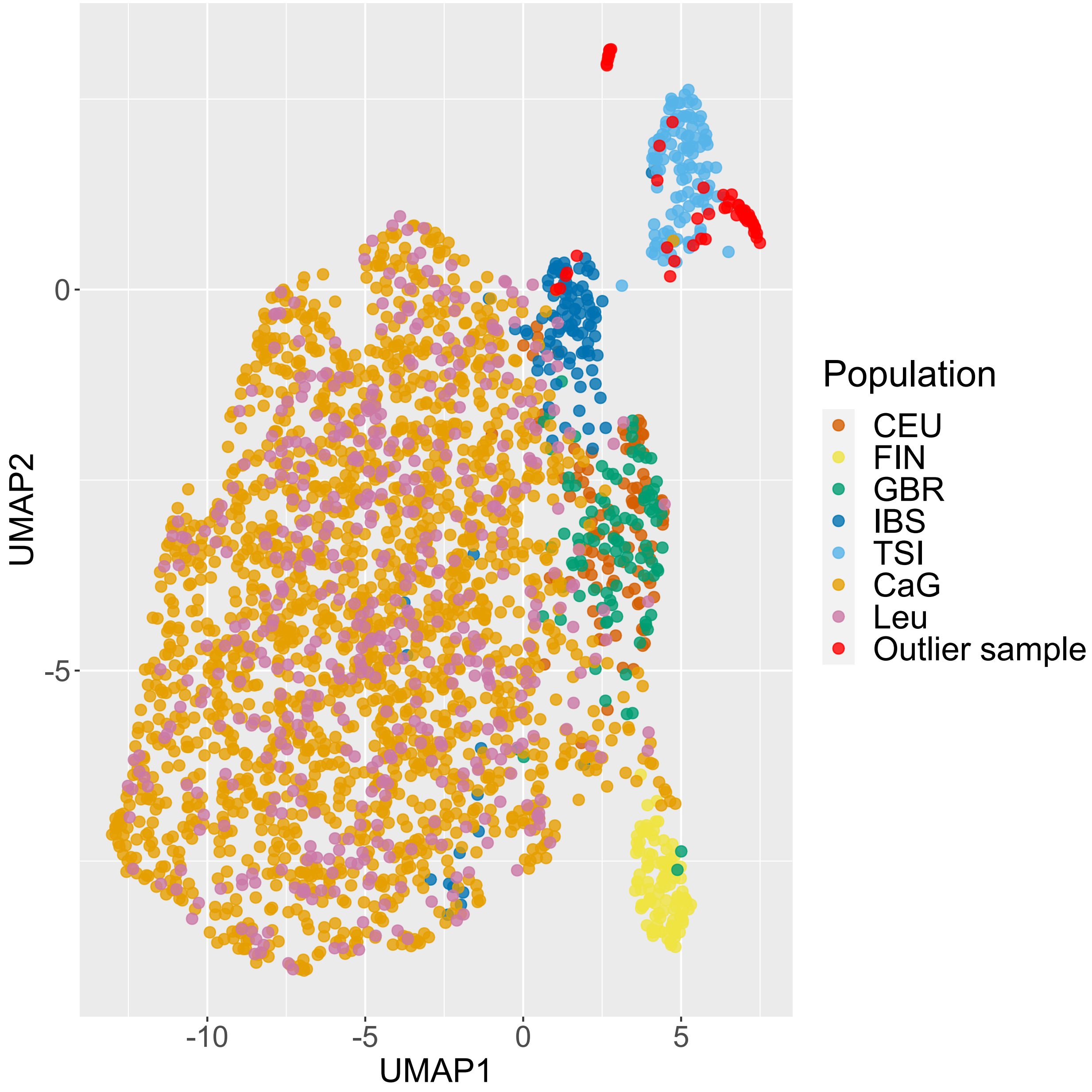

**Supplemental Figure 4.** Differentially expressed genes between Leucegene patients without mutations in *NPM1* (N=391) (**A**) and with mutations in *NPM1* (N=176) (**B**) with reference genotype (GG) and non-reference genotype (GA and AA) at rs3916765. Genes in red are HLA class II genes.

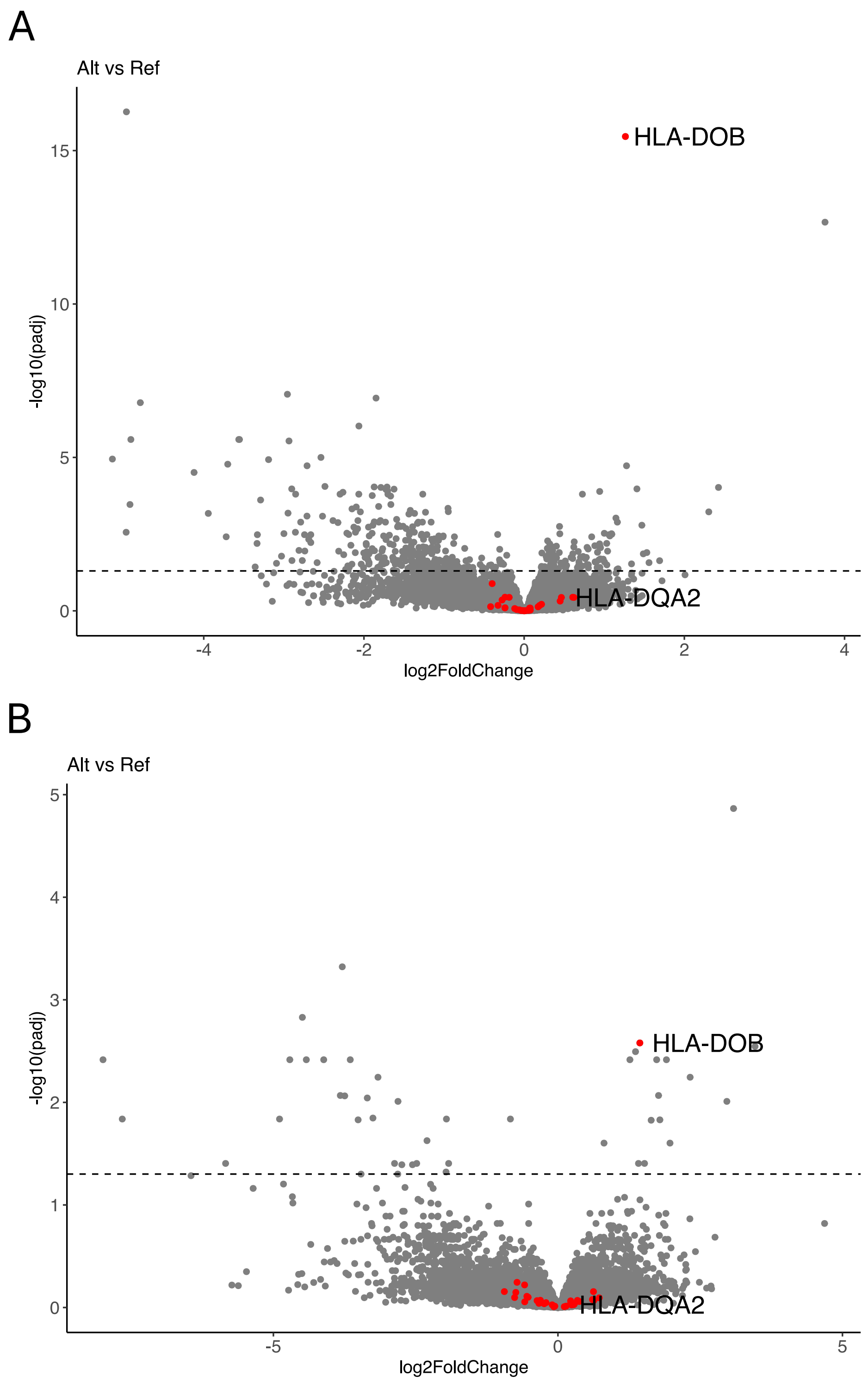

**Supplemental Figure 5.** Distributions of the polygenic risk scores (PRS) for leukemia and blood-cell traits in Leucegene AML cases and CARTaGENE (CaG) controls. The vertical lines denote the means of the score for the cases (in blue), and the controls (in red). ALL, acute lymphocytic leukemia; BASO, basophil count; CLL, chronic lymphocytic leukemia; EOS, eosinophil count; MONO, monocyte count; NEU, neutrophil count; PLT, platelet count; RBC, red blood cell count; WBC, total white blood cell count.

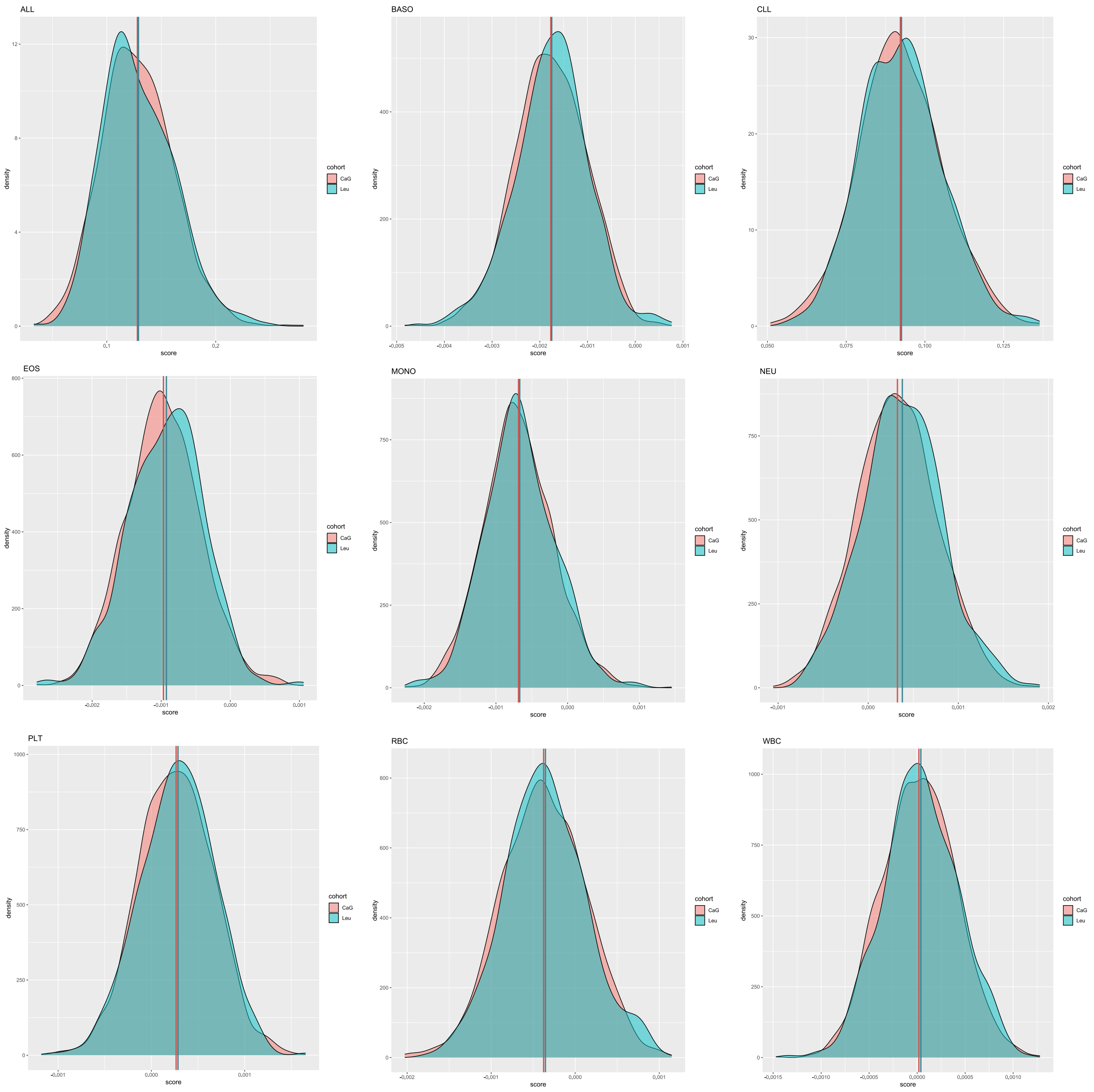

**Supplemental Figure 6.** Association of variants used to calculate the leukemia and blood-cell traits polygenic risk scores (PRS) with AML risk. Quantile-quantile plots (**A**,**C**) and Manhattan plots (**B**,**D**) of the association with AML risk in the Leucegene-CARTaGENE (CaG) dataset for the 2,032 variants used to calculate the PRS tested in this study (**Fig. 3 and Supplemental Fig. 5**). We extracted the association results from the GWAS with all Leucegene AML cases (n=567 cases and 1,865 controls) (**A**,**B**) or with cytogenetically-normal AML cases only (CN-AML) (n=239 cases and 1,865 controls) (**C**,**D**). Only two variants reach the statistical threshold after accounting for the number of tests performed (blue line, $\alpha$=0.05/2,032=2.4x10$^{-5}$) for all AML.

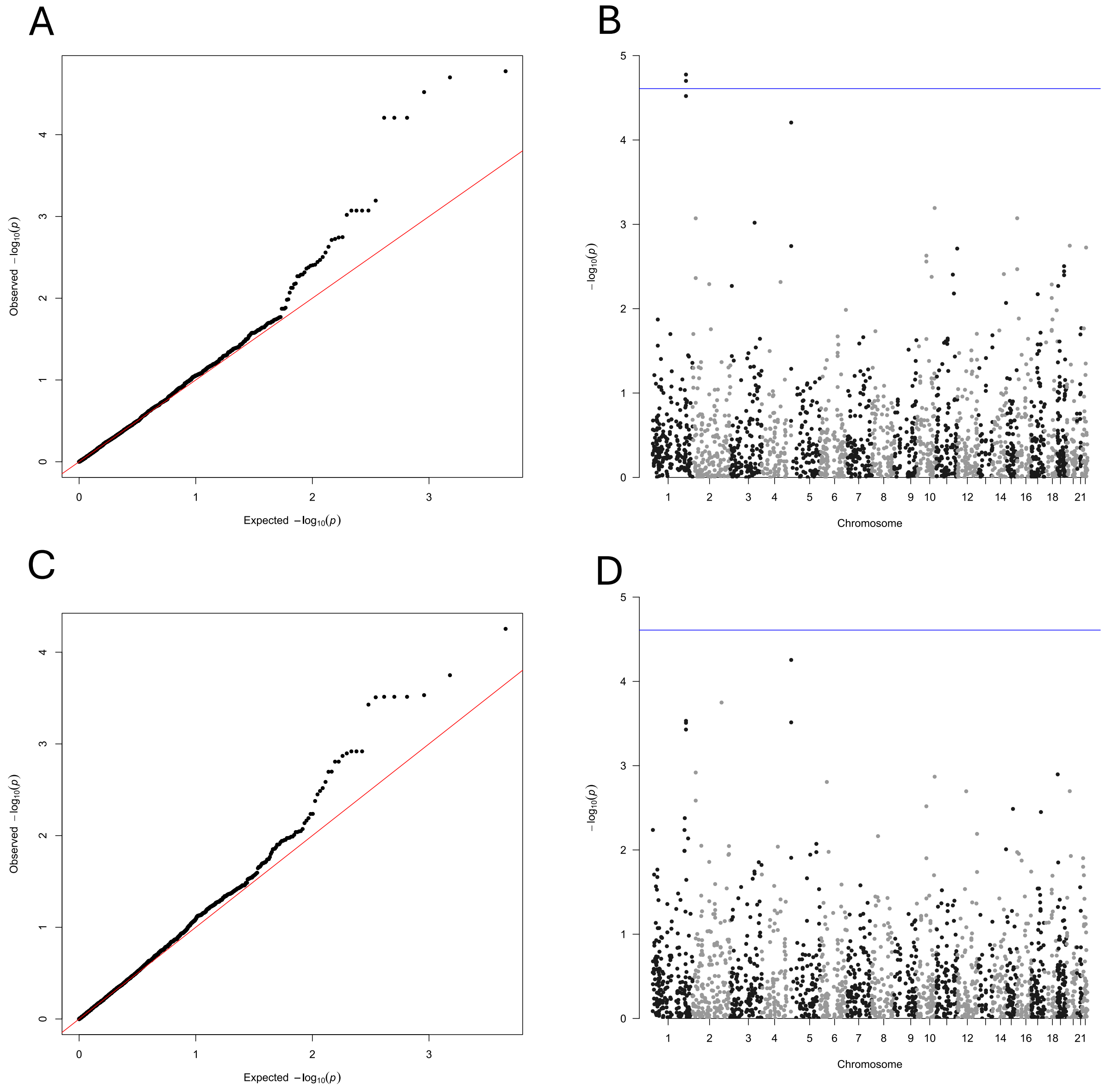

**Supplemental Table 1.** Replication results in the Leucegene-CARTaGENE dataset for two variants previously associated with acute myeloid leukemia (AML) by genome-wide association study (GWAS). These analyses are stratified using Leucegene AML cases with *NPM1* mutations (*NPM1*-mutated AML) or with *NPM1* mutations and a normal karyotype (CN-*NPM1*-mutated AML). OR [95% CI], odds ratio and 95% confidence interval; NFE, non-Finnish Europeans.

| | | | ***NPM1*-mutated-AML** N=177 cases, 1865 controls | | **CN-*NPM1*-mutated-AML** N=145 cases, 1865 controls | | **Allele frequency of the risk allele** | | | |
|---|---|---|---|---|---|---|---|---|---|---|
| **Variant** | **Risk allele** | **Gene** | **P-value** | **OR [95% CI]** | **P-value** | **OR [95% CI]** | **gnomAD NFE** | **Control** | **Cases *NPM1*-mutated AML** | **Cases CN-*NPM1*-mutated AML** |
| 6:32717773 G>A rs3916765 | G | *HLA-DQA* | 0.002 | 2.37 [1.38-4.08] | 0.002 | 2.70 [1.45-5.05] | 0.92 | 0.90 | 0.96 | 0.96 |
| 11:68164294 G>A rs4930561 | A | *KMT5B* | 0.11 | 1.20 [0.96-1.51] | 0.154 | 1.20 [0.93-1.53] | 0.52 | 0.49 | 0.54 | 0.54 |
